# Social Behaviors Associated with a Positive COVID-19 Test Result

**DOI:** 10.1101/2020.08.04.20168450

**Authors:** Sidra Speaker, Christine Doherty, Elizabeth R. Pfoh, Aaron Dunn, Bryan Hair, Victoria Shaker, Lynn Daboul, Michael B. Rothberg

## Abstract

**Objective:** To compare behaviors of individuals who tested positive for COVID-19 relative to non-infected individuals.

**Methods:** We sent COVID positive cases and age/gender matched controls a survey regarding their social behaviors via MyChart (online patient portal). We called cases if they did not complete the electronic survey within two days. Data was collected from May-June 2020. Survey responses for cases without a close contact and controls were compared using Pearson chi-square or Fisher’s Exact tests as appropriate.

**Results:** A total of 339 participants completed the survey (113 cases, 226 controls); 45 (40%) cases had known contact with COVID-19. Cases were more likely to have recently traveled (4% vs. 0%, p=0.01) or to work outside the home (40% vs. 25%, p=0.02). There was no difference in the rates of attending private or public gatherings, mask/glove use, hand-washing, cleaning surfaces and cleaning mail/groceries between cases and controls.

**Conclusions:** Sixty percent of cases had no known contact with COVID-19, indicating ongoing community transmission and underlining the importance of contact tracing. The greater percentage of cases who work outside the home provides further evidence for social distancing.

## Introduction

SARS-CoV-2 has infected >17 million individuals worldwide, with almost 700,000 deaths as of August 3, 2020(1). SARS-CoV-2 is highly transmissible, with an estimated reproductive number (R0) of 2, approximately double that of influenza(2). To mitigate the spread of COVID-19, the disease caused by SARS-CoV-2, there has been widespread promotion of public health measures. These measures, developed from experiences with other coronavirus outbreaks such as SARS, include isolation, quarantine, social distancing, and community containment. Isolation, the separation of symptomatic infected individuals from non-infected individuals, works best for diseases with a short incubation period. COVID-19 has a long incubation period and a substantial portion of transmission may occur from asymptomatic or pre-symptomatic individuals(3). Quarantine, restriction of movement of exposed individuals for the duration of the incubation period, is most useful when contacts of infected individuals can be readily traced. Social distancing involves limiting interactions among community members to decrease community spread. Finally, community containment involves restricting movements in entire geographical areas(4).

Population-based studies provide evidence that these measures have slowed the spread of COVID-19. In China, there was a significant reduction in cases following strict community containment(5). In four United States cities (New York City, Seattle, New Orleans, and San Francisco), new cases declined following the implementation of isolation, quarantining, and social distancing(6).

Less is known about the impact of specific recommendations, such as working from home, outdoor exercise, and disinfecting groceries/packages on the spread of the novel coronavirus. Hand-washing is one of the few recommendations to have been studied. A prospective cohort study from 2006-2009 found that individuals who engaged in hand-washing (6-10 times per day) had a reduced risk of acquiring coronavirus infections(7). In China, suboptimal hand hygiene was associated with COVID-19 infection among healthcare workers(8). It is important to identify which public health recommendations are most effective to support adherence and mitigate the psychosocial and economic impacts of unnecessary recommendations. In this case-control study, we describe patterns of COVID-19 prevention behavior among individuals in Ohio and Florida and differences between patients testing positive for COVID-19 versus uninfected patients.

## Materials and Methods

This work was approved by the Institutional Review Board of Cleveland Clinic. We surveyed adults with a COVID-19 PCR positive result from our institutional COVID-19 registry and gender- and age-matched controls from the Electronic Health Record (EHR) such that responses had approximately a 2:1 ratio. We sent all patients a 14-question survey regarding protective behaviors via MyChart, Cleveland Clinic’s online patient portal. We developed the survey by reviewing surveys of past epidemics(9,10) and the PhenX toolkit(11). Based on expert opinion and CDC guidelines, we included questions on contact with a person known to be infected with COVID-19, social distancing behaviors, and hygiene behaviors. The survey was pilot tested for clarity. Survey responses were stored in RedCap. We sent surveys to cases on the day of or the day following their first positive COVID-19 PCR test result, and to controls weekly. If cases did not complete the electronic survey within two days, we contacted them to administer it by phone. We excluded patients if they were non-English speaking, had dementia, resided in a skilled nursing facility, were currently hospitalized, or were not active on MyChart. Demographic data was extracted from the EHR from 5/12/2020 – 6/22/2020. After identifying individuals who reported close contact with a COVID positive individual, we compared the behavior of remaining cases and controls using Mann-Whitney U or Student’s t-tests (numeric variables) and Pearson chi-square or Fisher’s Exact tests (categorical variables) as appropriate. Statistical analyses were conducted using SAS (SAS Institute, Cary, NC) Studio v.9.4 and R Studio v.3.6.3.

Further information regarding the dataset is available from the corresponding author upon reasonable request.

## Results

A total of 113 cases and 226 controls completed the survey (response rate 9%), with 45 (40%) cases reporting having a close contact, compared to <1% of the controls. The median age was 54 and 63% were female.

A greater percentage of cases had someone in their residence who provided patient care (21% vs. 10%, p = 0.02). They were also more likely to live with someone experiencing upper respiratory tract symptoms within the past week (22% vs. 3%, p <0.001). Cases were more likely to work outside the home (40% vs. 25%, p = 0.02), to interact with 10+ people in their daily work (35% vs. 18%, p = 0.002), and to have traveled via airplane (4% vs. 0%, p =0.01) or RideShare/Taxi (4% vs. 0.4%, p = 0.04).

A greater percentage of cases were placed in isolation or quarantine (25% vs. 4%, p < 0.001). Cases were less likely to have gone outside to walk, hike, or exercise (52% vs. 88%, p<0.001).

There were no differences between cases and controls in the rates of attending gatherings, mask/glove use, hand-washing, cleaning surfaces and cleaning mail/groceries; and no difference in self-reported medical comorbidities.

**Table 1.**
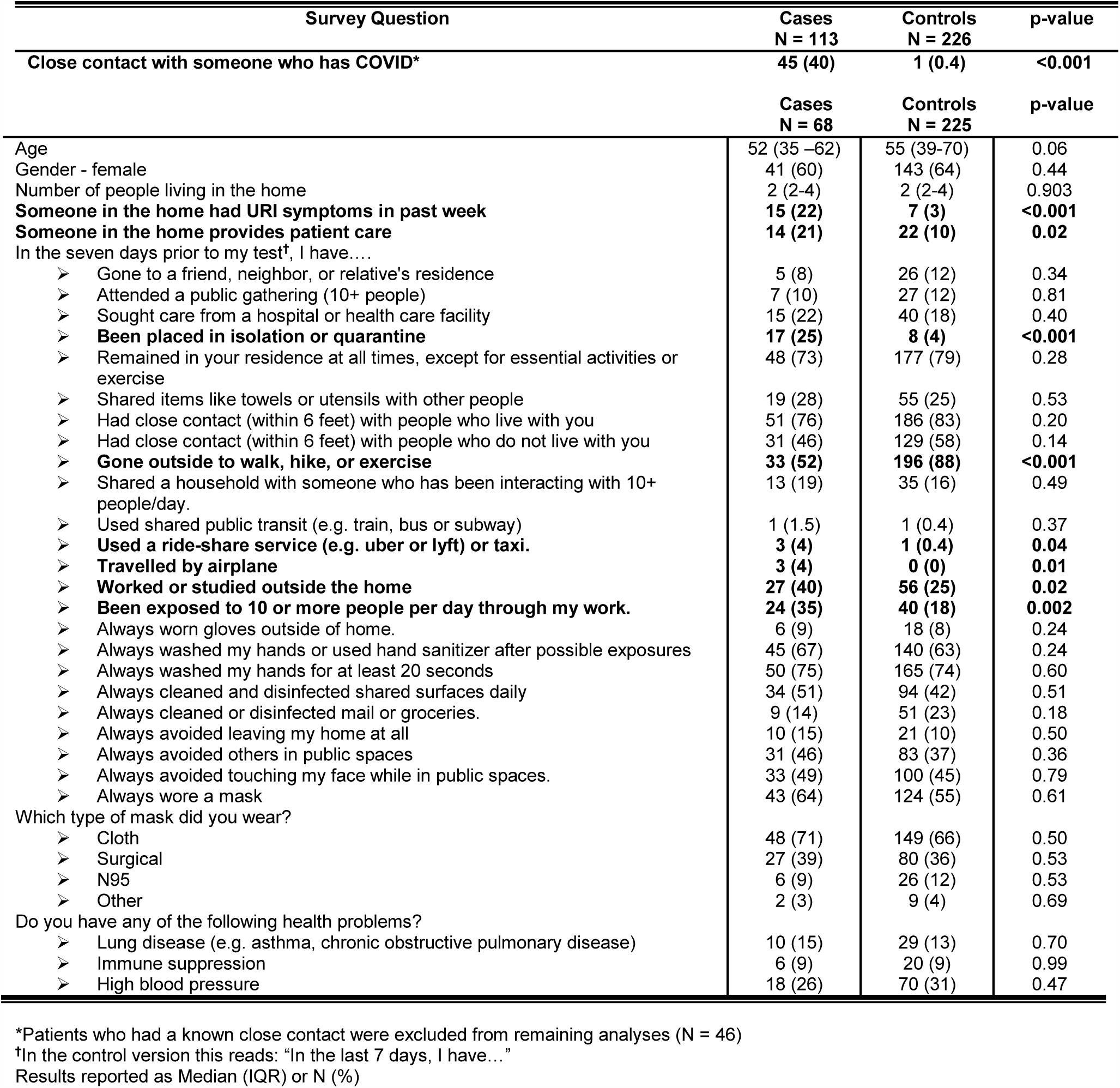
Survey Responses among Cases and Controls.

| Survey Question | Cases<br>N = 113 | Controls<br>N = 226 | p-value |
| --- | --- | --- | --- |
| Close contact with someone who has COVID* | 45 (40) | 1 (0.4) | <0.001 |
|  | Cases<br>N = 68 | Controls<br>N = 225 | p-value |
| Age | 52 (35 –62) | 55 (39-70) | 0.06 |
| Gender - female | 41 (60) | 143 (64) | 0.44 |
| Number of people living in the home | 2 (2-4) | 2 (2-4) | 0.903 |
| **Someone in the home had URI symptoms in past week** | **15 (22)** | **7 (3)** | **<0.001** |
| **Someone in the home provides patient care** | **14 (21)** | **22 (10)** | **0.02** |
| In the seven days prior to my test†, I have.... |  |  |  |
| ➤ Gone to a friend, neighbor, or relative's residence | 5 (8) | 26 (12) | 0.34 |
| ➤ Attended a public gathering (10+ people) | 7 (10) | 27 (12) | 0.81 |
| ➤ Sought care from a hospital or health care facility | 15 (22) | 40 (18) | 0.40 |
| ➤ **Been placed in isolation or quarantine** | **17 (25)** | **8 (4)** | **<0.001** |
| ➤ Remained in your residence at all times, except for essential activities or exercise | 48 (73) | 177 (79) | 0.28 |
| ➤ Shared items like towels or utensils with other people | 19 (28) | 55 (25) | 0.53 |
| ➤ Had close contact (within 6 feet) with people who live with you | 51 (76) | 186 (83) | 0.20 |
| ➤ Had close contact (within 6 feet) with people who do not live with you | 31 (46) | 129 (58) | 0.14 |
| ➤ **Gone outside to walk, hike, or exercise** | **33 (52)** | **196 (88)** | **<0.001** |
| ➤ Shared a household with someone who has been interacting with 10+ people/day. | 13 (19) | 35 (16) | 0.49 |
| ➤ Used shared public transit (e.g. train, bus or subway) | 1 (1.5) | 1 (0.4) | 0.37 |
| ➤ **Used a ride-share service (e.g. uber or lyft) or taxi.** | **3 (4)** | **1 (0.4)** | **0.04** |
| ➤ **Travelled by airplane** | **3 (4)** | **0 (0)** | **0.01** |
| ➤ **Worked or studied outside the home** | **27 (40)** | **56 (25)** | **0.02** |
| ➤ **Been exposed to 10 or more people per day through my work.** | **24 (35)** | **40 (18)** | **0.002** |
| ➤ Always worn gloves outside of home. | 6 (9) | 18 (8) | 0.24 |
| ➤ Always washed my hands or used hand sanitizer after possible exposures | 45 (67) | 140 (63) | 0.24 |
| ➤ Always washed my hands for at least 20 seconds | 50 (75) | 165 (74) | 0.60 |
| ➤ Always cleaned and disinfected shared surfaces daily | 34 (51) | 94 (42) | 0.51 |
| ➤ Always cleaned or disinfected mail or groceries. | 9 (14) | 51 (23) | 0.18 |
| ➤ Always avoided leaving my home at all | 10 (15) | 21 (10) | 0.50 |
| ➤ Always avoided others in public spaces | 31 (46) | 83 (37) | 0.36 |
| ➤ Always avoided touching my face while in public spaces. | 33 (49) | 100 (45) | 0.79 |
| ➤ Always wore a mask | 43 (64) | 124 (55) | 0.61 |
| Which type of mask did you wear? |  |  |  |
| ➤ Cloth | 48 (71) | 149 (66) | 0.50 |
| ➤ Surgical | 27 (39) | 80 (36) | 0.53 |
| ➤ N95 | 6 (9) | 26 (12) | 0.53 |
| ➤ Other | 2 (3) | 9 (4) | 0.69 |
| Do you have any of the following health problems? |  |  |  |
| ➤ Lung disease (e.g. asthma, chronic obstructive pulmonary disease) | 10 (15) | 29 (13) | 0.70 |
| ➤ Immune suppression | 6 (9) | 20 (9) | 0.99 |
| ➤ High blood pressure | 18 (26) | 70 (31) | 0.47 |

\*Patients who had a known close contact were excluded from remaining analyses (N = 46)
†In the control version this reads: "In the last 7 days, I have..."
Results reported as Median (IQR) or N (%)

## Discussion

In our study, only 40% of cases reported close contact with someone with COVID-19. Cases were more likely to report living with someone who had recent respiratory symptoms, possibly undetected infection. They were also more likely to provide direct patient care or live with someone who does. Health care workers are estimated to account for 3-19% of COVID-19 cases in the United States(12). These results reinforce the importance of testing and quarantining individuals with known or suspected exposure.

In our study, 60% of individuals reported no known contacts and were likely infected through community spread. One survey of infected patients found that most did not know the source of their infection, but over 80% worked outside the home in the two weeks preceding their diagnosis(13). Our study adds to this by comparing cases to uninfected controls, and confirming that work or study outside the home is a risk factor for SARS-CoV2 infection. In contrast, controls were more likely than cases to go outside to walk or exercise, suggesting that outdoor exercise is relatively safe.

Overall, compliance with recommended social distancing was suboptimal. At the time of survey administration there was low community spread in Ohio and Florida. About half the respondents had close contact with non-family members, only 60% always wore a mask, and two-thirds always washed their hands. At the same time, three-quarters remained in their homes except for essential activities, few attended gatherings of 10+ people and almost none travelled by public transportation. We found no difference between cases and controls in the rates of event attendance, mask or glove usage, hand-washing, and cleaning surfaces/mail/groceries. However, this does not prove that these interventions are not protective.

## Limitations

Given that our survey was computer and telephone based there may be non-response bias. Patients who responded may be more aware of and adherent to public health recommendations than the general public. Data were obtained via self-report and are subject to recall bias, however given the short time frame (within the past 7 days) this is less likely.

## Conclusions

Our study reveals the social behaviors and exposures of patients in Ohio and Florida from May – June 2020. The high percentage of infected individuals without known contacts or exposure to high-risk individuals reinforces the importance of contact tracing to reduce community spread. Infected individuals were more likely to have recently traveled and to work outside the home, emphasizing the importance of working remotely, social distancing, and minimizing travel.

## Acknowledgements

We would like to thank June Cassano and Gina Rupp for their assistance with the electronic survey deployment.

